# Identification of Somatic Structural Variants in Solid Tumors By Optical Genome Mapping

**DOI:** 10.1101/2021.02.04.21250683

**Authors:** David Y. Goldrich, Brandon LaBarge, Scott Chartrand, Lijun Zhang, Henry B. Sadowski, Yang Zhang, Khoa Pham, Hannah Way, Chi-Yu Jill Lai, Andy Wing Chun Pang, Benjamin Clifford, Alex R. Hastie, Mark Oldakowski, David Goldenberg, James R. Broach

**Affiliations:** Department of Otolaryngology – Head and Neck Surgery, Pennsylvania State University College of Medicine, Hershey, PA; Department of Biochemistry and Molecular Biology, Pennsylvania State University College of Medicine, Hershey, PA; Bionano Genomics, San Diego, CA

**Keywords:** optical genome mapping, solid tumors, cancer genomics

## Abstract

Genomic structural variants comprise a significant fraction of somatic mutations driving cancer onset and progression. However, such variants are not readily revealed by standard next generation sequencing. Optical genome mapping (OGM) surpasses short read sequencing in detecting large (>500bp) and complex structural variants (SVs) but requires isolation of ultra-high molecular weight DNA from the tissue of interest. We have successfully applied a protocol involving a paramagnetic nanobind disc to a wide range of solid tumors. Using as little as 6.5mg of input tumor tissue, we show successful extraction of high molecular weight genomic DNA that provides a high genomic map rate and effective coverage by optical mapping. We demonstrate the system’s utility at identifying somatic SVs affecting functional and cancer-related genes for each sample. Duplicate/triplicate analysis of select samples shows intra-sample reliability but also intra-sample heterogeneity. We also demonstrate that simply filtering SVs based on a GRCh38 human control database provides high positive and negative predictive values for true somatic variants. Our results indicate that the solid tissue DNA extraction protocol, OGM and SV analysis can be applied to a wide variety of solid tumors to capture SVs across the entire genome with functional importance in cancer prognosis and treatment.

## Introduction

One of the hallmarks of cancer is genomic instability, which often affects genes controlling cell division and genome integrity. The resulting alterations include single nucleotide variant (SNV) point mutations as well as structural variants (SVs), in which larger DNA segments undergo chromosomal perturbations such as deletions, insertions, duplications, inversions, and translocations. For instance, recurrent translocations, such as the Philadelphia chromosome, can activate oncogenes but at the same time reveal avenues for implementing or developing effective targeted drug therapies [1-4]. Likewise, SV identification plays an increasingly important role in cancer diagnosis and prognosis [5,6], and SVs have been shown to play a crucial role in intra-tumoral genetic heterogeneity [7]. Therefore, SV identification and analysis is important to understanding oncogenesis and tumor behavior.

Short read sequencing can readily detect many SNVs, but is less successful in detecting SVs, by either alignment-based or assembly-based methods [8]. Since alignment-based approaches rely on mapping reads to unique positions, repetitive and low-complexity genomic regions can lead to misalignment and false positive SV calls. Additionally, homologous alleles may be incorrectly combined, leading to haploid assembly only representing a single allele or chimeric assemblies mixing alleles. Whole genome and cytogenetic approaches such as whole genome sequencing (WGS), karyotyping, fluorescent in-situ hybridization (FISH) and CNV microarrays also contain significant limitations. Karyotyping provides a comprehensive view of the entire genome but carries limited resolution of ∼5Mb and in most cases requires culturing cells before preparing chromosomes. FISH has higher resolution but requires prior knowledge as to which loci to test and has limited throughput. CNV-microarrays offers resolution down to multiple Kb but are insensitive to balanced chromosomal aberrations such as translocations and inversions, are unable to detect low-frequency allelic changes, and cannot distinguish tandem duplications from insertions in trans. Finally, WGS has difficulty with de-novo genome assembly and resolving duplications and repeated sequences [8-10]. Therefore, alternative methods are required to preserve long-range genomic structural information.

Optical genome mapping (OGM) has emerged as a viable option for analyzing large genomes for SVs. OGM preserves long-range information by imaging entire intact molecules of DNA in their native state and, as a result, has contributed to constructing reference genome assemblies, including those for maize, mouse, goat, and humans [11-28]. OGM can detect large (>500bp) and complex SVs that are difficult to detect using traditional short read sequencing alone. OGM preparation and analysis workflow has been successfully applied to liquid-phase tumor and cell culture SV analyses. For instance, Neveling et al. successfully analyzed SV and CNV calls and identified novel potential drivers of leukemia using bone marrow and blood samples [29]. Xu et al utilized OGM to detect previously unrecognized SVs in leukemia patients implicated in oncogenesis and patients’ survival [5]. Chan et al used OGM to visualize complex gene fusion maps in previously studied liposarcoma cell line [30]. Pang et al demonstrated the ability of OGM to identify novel somatic SVs in melanoma and other well-studied cancer cell lines [31]. Dixon et al integrated OGM to high-throughput chromosome conformation capture (Hi-C) and WGS to systematically detect SVs in leukemia and other cell lines [32].

Despite its success in visualizing SVs in liquid tumors, OGM has not yet seen widespread application in solid tissue tumors, due primarily to the difficulty of obtaining high quality, high molecular weight DNA from solid tumor samples. Nonetheless, previous work has shown the feasibility of high-quality high molecular weight DNA isolation and analysis using earlier workflow iterations [33], and recent feasibility studies have shown the importance of OGM application to solid tumor analysis [7,34,35]. Peng et al demonstrated large SVs not detected by WGS implicated in metastatic lung squamous cell carcinoma [7], and Jaratlerdiri et al and Crumbaker et al similarly found SVs impacting oncogenic and tumor suppressing genes not identified by NGS or WGS alone in prostate cancer [34,35]. However, these previous methods for extracting gDNA from solid tissue were either prohibitively expensive or yielded low quantities of DNA [36]. We demonstrate here the successful implementation of a workflow to generate ultrahigh molecular weight gDNA and subsequent SV analysis for 20 solid tumor samples comprising a wide variety of solid tissue organ systems.

## Materials and Methods

### Tumor Samples

Solid tissue was collected following surgical resection for 10 tumors: four squamous cell carcinomas of the tongue, three anaplastic carcinomas of the thyroid, one liver hepatocellular carcinoma, one lung pleomorphic carcinoma, and one bladder tumor. Patients were consented under protocols approved by the Penn State Health Institution Review Board. Tissue was flash frozen and stored at −80°C in the Penn State Institute for Personalized Medicine (IPM). Ten additional fresh frozen solid tumor samples were acquired from BioIVT for the following tumor types: lung adenosquamous carcinoma, liver hepatocellular carcinoma, bladder papillary urothelial carcinoma, kidney renal cell carcinoma, breast ductal carcinoma in-situ, prostate invasive adenocarcinoma, brain anaplastic astrocytoma, ovarian serous carcinoma, colon adenocarcinoma, and papillary thyroid carcinoma.

### Bionano Optical Genome Mapping

#### Ultra-High Molecular Weight gDNA Isolation from Solid Tissue

(Figure 1). A 10mg section of frozen tissue was sliced from its parent piece on a sterilized aluminum block over dry ice. The tissue was minced briefly into 1-2mm pieces with a scalpel, and placed into a 15ml conical tube containing 2ml of chilled Bionano Prep SP Animal Tissue Homogenization Buffer (HB) on ice. The tube was then placed in a beaker of ice for effective cooling of the sample during subsequent blending procedure. The tissue was blended at full speed for 10 seconds using the Tissueruptor II (Qiagen). Following tissue disruption, 6ml of additional chilled HB was added to the sample, and the resultant 8ml was filtered through a 40µm nylon cell filter within a 50ml conical tube.

**Figure 1.**
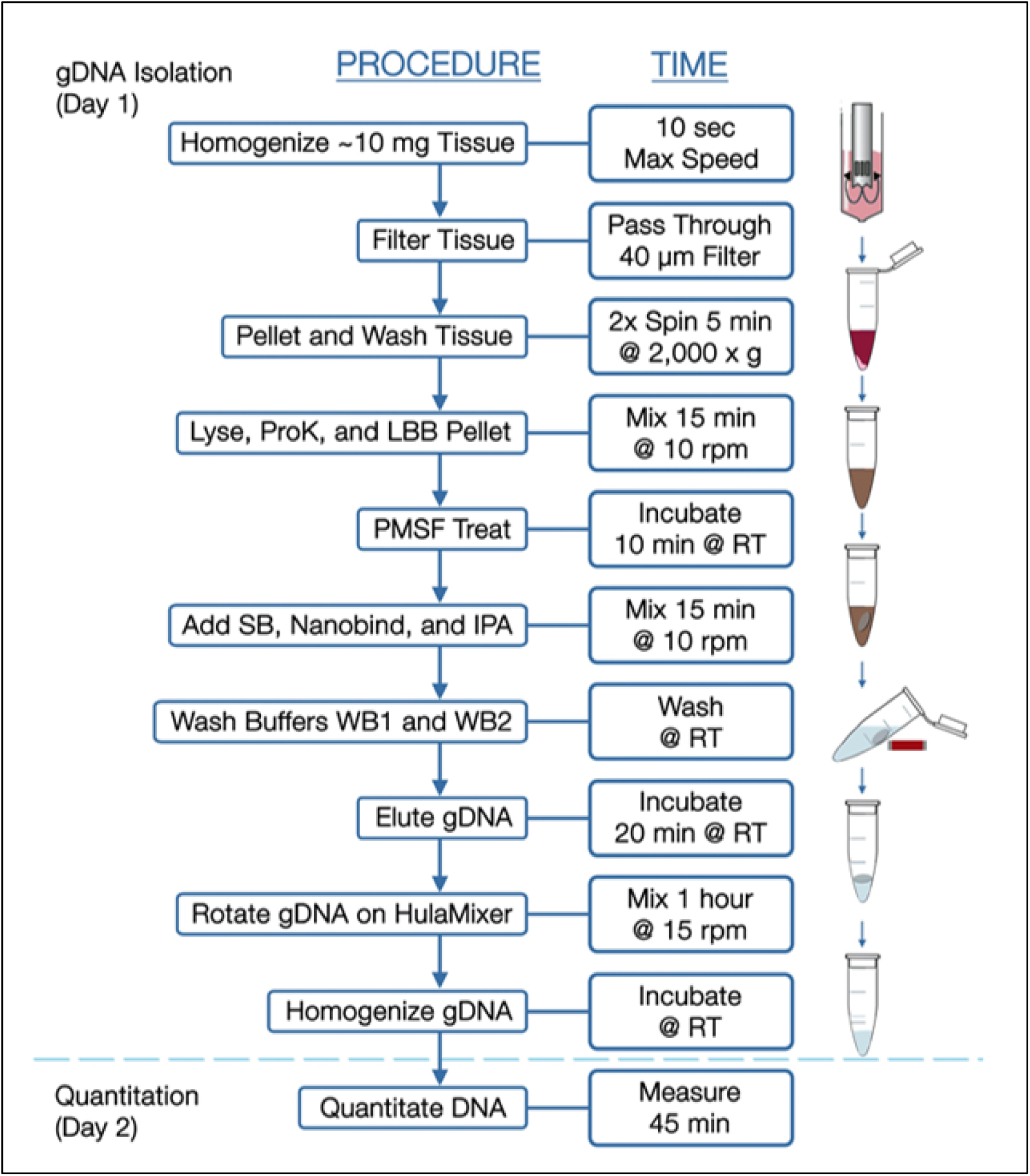
Workflow for Isolation of High Molecular Weight DNA from Solid Tumors. See text for detailed description of the workflow.

Homogenate was transferred to a new 15ml tube, inverted, centrifuged at 2000g for 5 minutes at 4°C using a swinging bucket rotor, and decanted. The cell pellet was then resuspended in 300μl of Wash Buffer A (Bionano) and transferred to a 1.5ml Protein LoBind tube. 700μl additional Wash Buffer A was added to the 15ml tube, pipette mixed, and transferred to the same 1.5ml tube, followed by centrifuge at 2000g for 5 minutes at 4°C. The supernatant was then decanted and the pellet was resuspended in residual volume. 50μl Proteinase K (Bionano) was added to the sample and incubated at room temperature for 3 minutes. 225μl Buffer Lysis and Binding Buffer (LBB, Bionano) was then added, and samples were inverted 15 times to mix. After HulaMixer rotation at 10rpm for 15 minutes at room temperature, 10μl of 100mM Phenylmethylsulfonyl Fluoride Solution (PMSF, Sigma-Aldrich) was added to inactivate Proteinase K, followed by 85μl of Salting Buffer (SB, Bionano).

A single 4mm paramagnetic Nanobind Disc (Bionano) was transferred to the lysate, in addition to 400μl 100% isopropanol. Binding of gDNA to the disc was facilitated by HulaMixer rotation for 15 minutes at room temperature at 10 RPM. Samples were then placed into a clear Dynamag tube rack for quick inversions to further increase gDNA binding. The rack was then inverted and joined to the Dynamag magnetic base. The apparatus was slowly brought upright to allow the disc to adhere to the magnet. The supernatant was removed and 700μl Wash Buffer 1 (Bionano) was dispensed onto the disc, followed by four inversions of the clear rack for washing. This washing step was performed twice and followed by two washes of 500μl Wash Buffer 2 (Bionano). After final decanting, the disc was transferred to a new 1.5 ml Protein LoBind tube using the Magnetic Retriever (Bionano). Elution Buffer (65μl, Bionano) was added and the sample was incubated for 20 minutes at room temperature. gDNA was then transferred to a 2.0ml microfuge tube. The previous tube containing the disc was spun on a benchtop microcentrifuge for 5 seconds and the residual eluate transferred to the 2.0 ml tube. The gDNA was slowly pipetted to homogenize and then rotated via HulaMixer at room temperature for 1 hour at 15 RPM. The sample was then pulse spun and allowed to equilibrate overnight at room temperature.

#### Ultra-High Molecular Weight gDNA Isolation from Blood

A 650μl aliquot of frozen blood was thawed to room temperature. The sample was inverted 10 times to mix and then pulse spun. 20μl was dispensed onto a Hemocue cuvette and cell density was measured. The amount of blood necessary to obtain 1.5 × 10^6^ cells was calculated (typically 200-600μl). Sample was again inverted 10 times and briefly pulse spun before the calculated volume was transferred to a 1.5ml Protein LoBind tube.

The resultant blood aliquot was spun for 2 minutes at 2,200 x g at room temperature to obtain a WBC pellet. After removing the supernatant, 40μl Stabilizing Buffer and 50μl Proteinase K (Bionano) were added the WBC pellet. After resuspension, the sample was incubated at room temperature for 3 minutes. 225μl Buffer LBB (Bionano) was added to the sample and mixed by inversion fifteen times. The sample was pulse spun and 10μl of 100mM PMSF (Sigma-Aldrich) was added to the sample, followed by capping and inversion five times and a brief pulse spin. The sample was incubated at room temperature for 10 minutes. Isolation of DNA was then performed essentially as described above for tumor tissue.

#### Direct Label and Staining (DLS)

750ng of gDNA, determined by Broad Range dsDNA Qubit Assay, was mixed with Direct Label Enzyme-1 (DLE-1) Buffer (Bionano Genomics, Part#20350), DL-Green (Bionano Genomics, Part#20352), and DLE-1 Enzyme (Bionano Genomics, Part#20351) and incubated for 2 hours at 37°C in a thermocycler. 5µl of Proteinase K solution was then added to the reaction and incubated for 30 minutes at 50°C. During Proteinase K digestion, a DLS membrane (Bionano Genomics, Part#20358) was placed hydrated with 25μL of DLE-1 buffer in one well of a DLS-microplate (Bionano Genomics, Part#20357). The labeled DNA was transferred onto this membrane, incubated at room temperature for one hour, transferred onto another membrane with 25μL DLE-1 buffer and incubated for 30 minutes at room temperature. Sample exposure to light was avoided during these steps. Following this drop dialysis cleanup, 5x DTT (Bionano Genomics, Part#20354), Flow Buffer (Bionano Genomics, Part#20353), DNA stain (Bionano Genomics, Part#20356), and nuclease free water were added to the DNA in a 2ml amber tube and the tube was mixed in a hula mixer at 5rpm for 2 hours at room temperature. The sample was then spun down and stored overnight at room temperature protected from light. The DNA concentration of each labeled sample was confirmed within 4-12 ng/µl by High Sensitivity dsDNA Qubit Assay and then loaded onto a Bionano Saphyr® Chip (Bionano Genomics, Part#20319) and run on the Bionano Saphyr® instrument, targeting approximately 300x human genome coverage.

### Bionano Access and Solve Pipeline

Genome analysis was performed using Rare Variant Analysis in Bionano Access 1.6 and Bionano Solve 3.6, which captures somatic SVs occurring at low allelic fractions. Briefly, molecules of a given sample dataset were first aligned against the public Genome Reference Consortium GRCh38 human assembly. SVs were identified based on discrepant alignment between sample molecules and GRCh38, with no assumptions about ploidy. Consensus genome maps (*.cmaps) were then assembled from clustered sets of at least three molecules that identify the same variant. Finally, the genome maps were realigned to GRCh38, with SV data confirmed by consensus forming final SV calls. SVs were then annotated with known canonical gene set present in GRCh38, as well as estimated population frequency for each structural variant detected by comparing to a custom control database (n=297) from Bionano Genomics.

### Data Comparison

Whole genome imaging data were compared to the human reference genome GRCh38 (hg38) to retain only those SVs not present in the reference genome. SVs were further filtered to eliminate any variant observed in any of the Bionano control samples or, if available, patient-matched blood. Bionano Access-created csv files containing filtered SVs were analyzed to compare SV content across samples. For tissue samples with associated blood samples, control database filtration efficacy was compared to blood-filtering efficacy at identification of somatic mutations. For duplicate/triplicate samples, filtered SVs were compared to determine intra-sample reliability. For identification of cancer related genes, the set of genes affected by SVs in each of the samples was compared to the list of genes causally implicated in cancer available in the Cosmic Cancer Gene Census database (v92) [37] (https://cancer.sanger.ac.uk/census).

## Results

### Patient Clinical Characteristics

Clinical data for the patients from whom tumor samples were acquired are shown in Table 1. 60% (12/20) patients were male, with mean age of 73.5 years at sample acquisition. 45% (9/20) patients identified as Caucasian, 40% (8/20) as Asian, 5% (1/20) as Hispanic and 10% (2/20) not identifying. The majority of IPM-sourced tumor samples were obtained from Caucasian patients (7/10) while the majority of the BioIVT-sourced tumor samples were obtained from patients of Asian ethnicity (8/10). In terms of overall risk factors, 55% (11/20) of patients were self-described current or former tobacco users and 45% (9/20) endorsed some history of alcohol use.

**Table 1.**
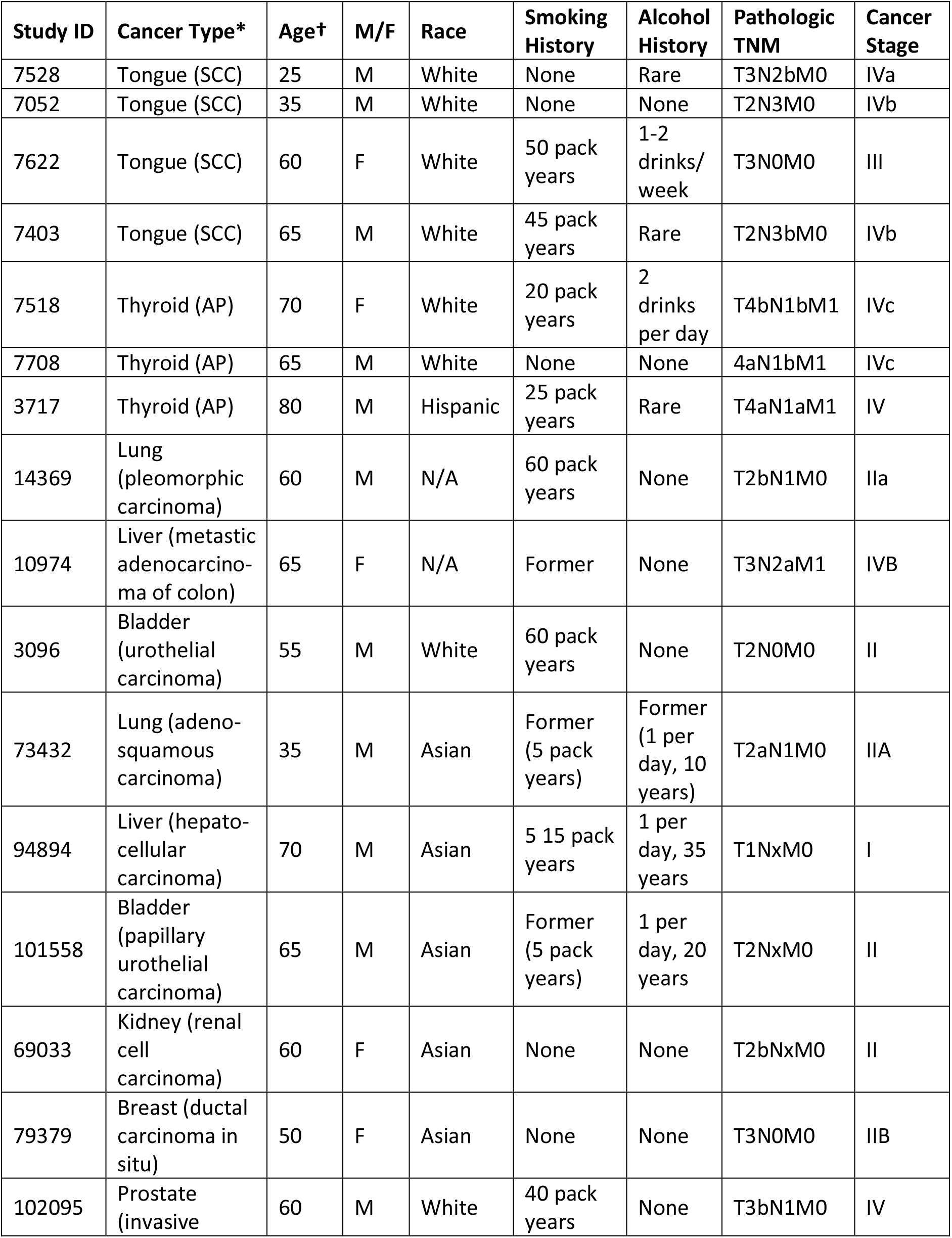
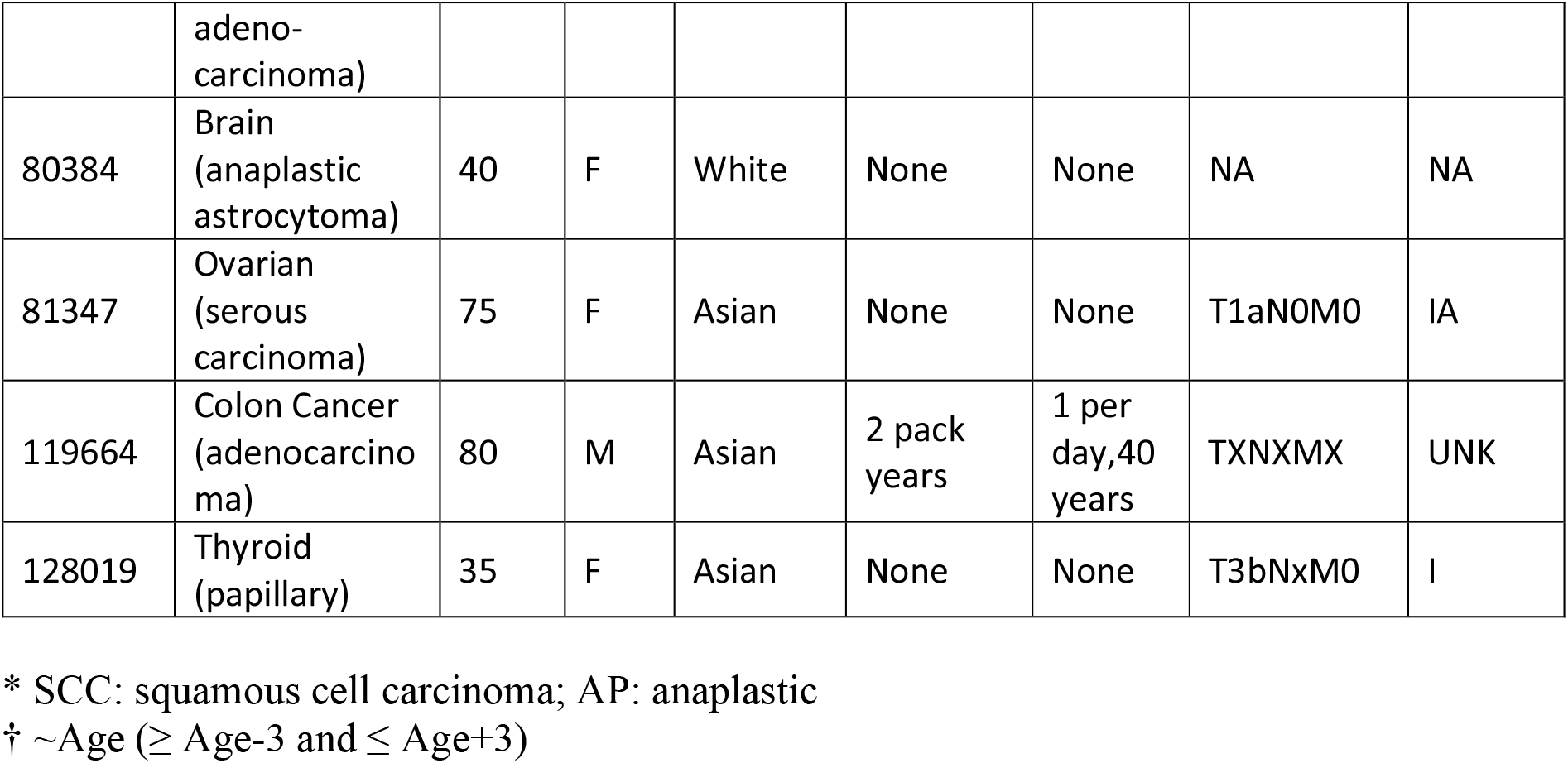
Patient Demographics and Tumor Characteristics.

| Study ID | Cancer Type* | Age† | M/F | Race | Smoking History | Alcohol History | Pathologic TNM | Cancer Stage |
| --- | --- | --- | --- | --- | --- | --- | --- | --- |
| 7528 | Tongue (SCC) | 25 | M | White | None | Rare | T3N2bM0 | IVa |
| 7052 | Tongue (SCC) | 35 | M | White | None | None | T2N3M0 | IVb |
| 7622 | Tongue (SCC) | 60 | F | White | 50 pack years | 1-2 drinks/ week | T3N0M0 | III |
| 7403 | Tongue (SCC) | 65 | M | White | 45 pack years | Rare | T2N3bM0 | IVb |
| 7518 | Thyroid (AP) | 70 | F | White | 20 pack years | 2 drinks per day | T4bN1bM1 | IVc |
| 7708 | Thyroid (AP) | 65 | M | White | None | None | 4aN1bM1 | IVc |
| 3717 | Thyroid (AP) | 80 | M | Hispanic | 25 pack years | Rare | T4aN1aM1 | IV |
| 14369 | Lung (pleomorphic carcinoma) | 60 | M | N/A | 60 pack years | None | T2bN1M0 | Ila |
| 10974 | Liver (metastatic adenocarcinoma of colon) | 65 | F | N/A | Former | None | T3N2aM1 | IVB |
| 3096 | Bladder (urothelial carcinoma) | 55 | M | White | 60 pack years | None | T2N0M0 | II |
| 73432 | Lung (adenosquamous carcinoma) | 35 | M | Asian | Former (5 pack years) | Former (1 per day, 10 years) | T2aN1M0 | IIA |
| 94894 | Liver (hepatocellular carcinoma) | 70 | M | Asian | 5 15 pack years | 1 per day, 35 years | T1NxM0 | I |
| 101558 | Bladder (papillary urothelial carcinoma) | 65 | M | Asian | Former (5 pack years) | 1 per day, 20 years | T2NxM0 | II |
| 69033 | Kidney (renal cell carcinoma) | 60 | F | Asian | None | None | T2bNxM0 | II |
| 79379 | Breast (ductal carcinoma in situ) | 50 | F | Asian | None | None | T3N0M0 | IIB |
| 102095 | Prostate (invasive) | 60 | M | White | 40 pack years | None | T3bN1M0 | IV |
|  | adeno-<br>carcinoma) |  |  |  |  |  |  |  |
| 80384 | Brain<br>(anaplastic<br>astrocytoma) | 40 | F | White | None | None | NA | NA |
| 81347 | Ovarian<br>(serous<br>carcinoma) | 75 | F | Asian | None | None | T1aN0M0 | IA |
| 119664 | Colon Cancer<br>(adenocarcino<br>ma) | 80 | M | Asian | 2 pack<br>years | 1 per<br>day,40<br>years | TXNXMX | UNK |
| 128019 | Thyroid<br>(papillary) | 35 | F | Asian | None | None | T3bNxM0 | I |

\* SCC: squamous cell carcinoma; AP: anaplastic
† ~Age ( ≥ Age-3 and ≤ Age+3)

The tumor samples consisted of a variety of stages (Table 1). 75% (3/4) of tongue cancer samples and 100% (3/3) anaplastic thyroid cancers were stage IV cancers, while 100% (2/2) lung and (2/2) bladder cancers were stage II. Limited tumor data was available for the commercially available BioIVT-sourced tumor samples.

#### DNA quality metrics

All 20 solid tumors yielded high molecular weight gDNA (Table 2). The average concentration across all samples following gDNA isolation was 120 ng/µl by Broad Range dsDNA Qubit Assay. All eluted gDNA were well above the minimal concentration required for DLS labeling (35ng/µl) and the average final DNA yields for each tumor ranged from 1.2-16.4µg/10mg input tissue. Analysis on a Saphyr instrument following DLS labeling revealed that samples achieved an average label density of 14.4/100Kbp, average filtered N50 (>20kbp) DNA size of 242Kbp, average filtered N50 (>150kbp) DNA size of 315Kbp, map rate of 82.62%, effective reference coverage of 320x and average effective DNA throughput (≥150Kbp) of 50Gbp/scan. Rare Variant Pipeline Analysis of the samples yielded an average of 82.4% of molecules aligning to the reference genome. These values are all well above the acceptable range for obtaining high quality data and none of the samples failed any of these quality control metrics.

**Table 2.** Single Molecule Quality Report Metrics.

| **Tissue** | **#** | **Input**<br>**(mg)** | **DNA**<br>**(ng/μl)** | **DNA Yield**<br>**(μg/mg)** | **N50 Kbp**<br>**(>150Kbp)** | **Labels/**<br>**100kbp** | **Map**<br>**Rate**<br>**(%)** | **Gbp**<br>**/scan** | **Effective**<br>**Coverage** |
| --- | --- | --- | --- | --- | --- | --- | --- | --- | --- |
| 7528 (tongue) | 1 | 17.5 | 37 | 0.12 | 317 | 12.3 | 58.8 | 53 | 237x |
| 7052 (tongue) | 1 | 17.1 | 81 | 0.28 | 287 | 15.2 | 82.4 | 37 | 345x |
| 7622 (tongue) | 1 | 18.7 | 160 | 0.51 | 361 | 13.4 | 75.8 | 64 | 317x |
| 7403 (tongue) | 1 | 18 | 79 | 0.26 | 272 | 14.8 | 72.8 | 33 | 304x |
| 7518 (thyroid) | 1 | 8.6 | 28 | 0.20 | 265 | 14.4 | 76.6 | 26 | 312x |
| 7708 (thyroid) | 1 | 10.6 | 85 | 0.47 | 356 | 13.1 | 61.2 | 35 | 253x |
| 3717 (thyroid) | 1 | 13.2 | 49 | 0.22 | 320 | 14.5 | 88.2 | 58 | 371x |
| 14369 (lung) | 1 | 11.4 | 87 | 0.45 | 323 | 14.0 | 89.6 | 36 | 372x |
| 10974 (liver) | 1 | 6.5 | 82 | 0.74 | 289 | 15.2 | 87.9 | 49 | 360x |
| 3096 (bladder) | 1 | 9.4 | 59 | 0.37 | 319 | 13.8 | 78.3 | 39 | 325x |
| 73432 (lung) | 3 | 9.6 | 128 | 0.86 | 304 | 15.0 | 90.4 | 51 | 339x |
| 94894 (liver) | 2 | 9.0 | 196 | 1.41 | 306 | 14.9 | 89.3 | 84 | 325x |
| 101558<br>(bladder) | 3 | 9.7 | 245 | 1.64 | 357 | 15.2 | 91.8 | 66 | 338x |
| 69033 (kidney) | 3 | 10 | 96 | 0.63 | 269 | 14.6 | 83.5 | 41 | 296x |
| 79379 (breast) | 3 | 13.3 | 183 | 1.04 | 395 | 14.2 | 84.1 | 77 | 288x |
| 102095<br>(prostate) | 3 | 10.3 | 113 | 0.72 | 361 | 14.8 | 85.1 | 62 | 295x |
| 80384 (brain) | 2 | 10.5 | 168 | 1.06 | 292 | 14.6 | 90.2 | 42 | 306x |
| 81347 (ovary) | 2 | 10.5 | 168 | 1.05 | 292 | 14.6 | 90.2 | 42 | 330x |
| 119664 (colon) | 2 | 11.3 | 231 | 1.33 | 330 | 14.9 | 88.6 | 42 | 274x |
| 128019<br>(thyroid) | 4 | 10 | 126 | 0.77 | 294 | 14.5 | 87.6 | 64 | 294x |
| Average | 1.9 | 11.8 | 120. | 0.71 | 315. | 14.4 | 82.6 | 50.0 | 314x |

Average values are presented for samples with multiple replicates

#### Identification of somatic structural variants

Rare Variant analysis of the samples revealed a large numbers of variants in each sample, only a fraction of which were likely somatic (Figure 2). The unfiltered analysis yielded an average of 1633 total SVs per sample (range 1241-2000), which include both somatic and germline polymorphic variants. These consisted predominantly of insertions and deletions, with an average of 712 insertions and 604 deletions, a fewer number of inversion (an average of 153) and duplications (an average of 123), and relatively few translocations (an average of 41). Eliminating those SVs found in Bionano’s control database of known polymorphic SVs reduced the number of putative somatic structural mutations by 91% to an average of 124 total SVs per sample (Figure 2). Most of the variants eliminated were insertions and deletions, of which on average 97% and 94%, respectively, were removed. On the other hand, less than 0.2% of the translocations were flagged as polymorphic, consistent with the fact that almost no translocations persist in the population as polymorphism.

**Figure 2.**
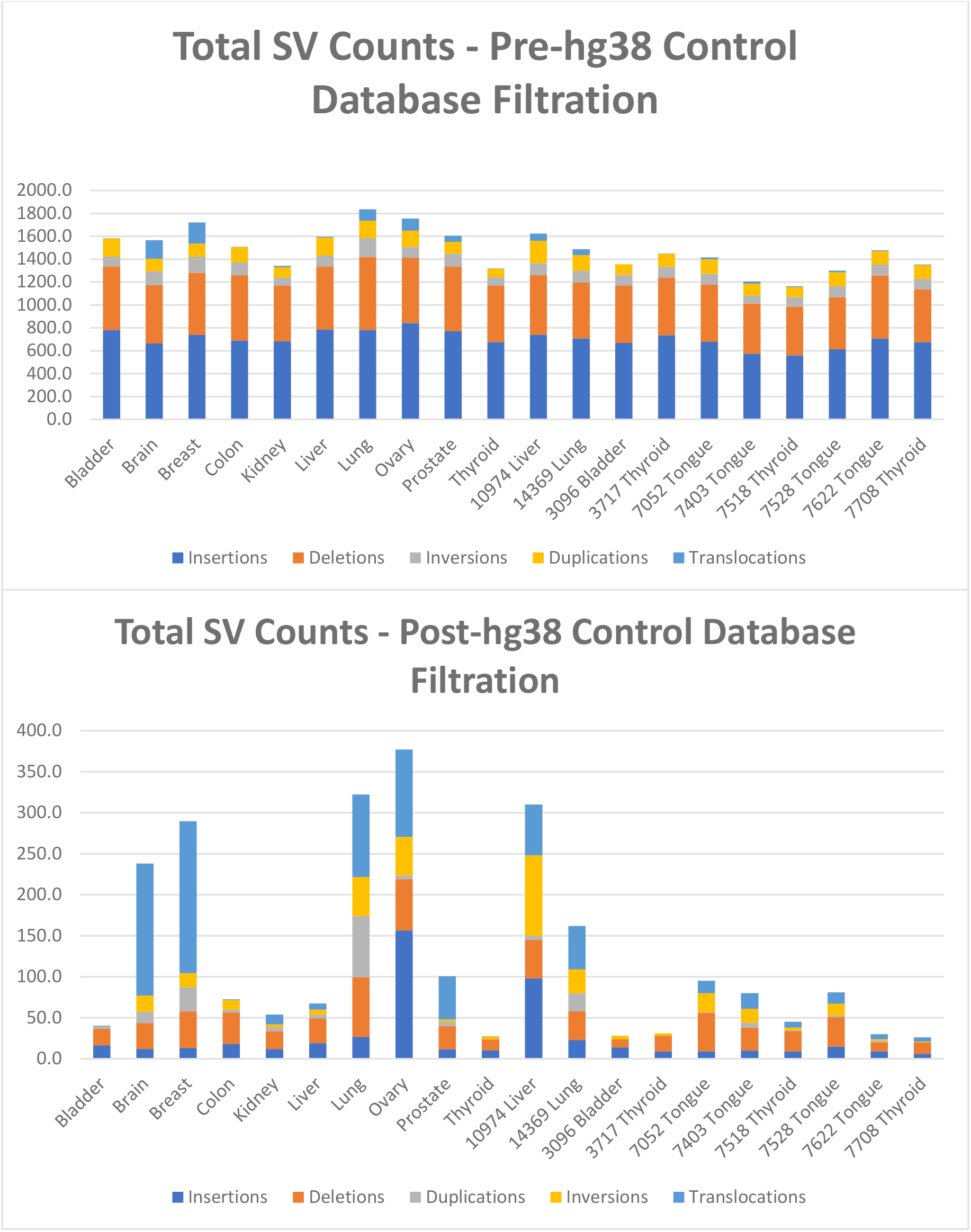
Total and Somatic Structural Variants Present in Tumor Samples. Upper panel: SV counts as determined using the Bionano Rare Variant pipeline, before control database filtration. SV counts are averages for duplicate and triplicate samples. Lower panel: SV counts after filtering total SVs to remove known polymorphic SV found in Bionano’s GRCh38 control database. SV counts are averages for duplicate and triplicate samples.

To determine the efficacy of identifying somatic SVs by filtering against Bionano’s database of known polymorphisms, we used as a gold standard the blood samples from four patients from whom we had obtained tongue tumors. That is, we determined the true somatic mutations in each of these four tumors by eliminating those SVs identified in each of the tumors that were also present in the corresponding blood sample. We could then compare those true somatic variants to the list of somatic variants predicted by filtering against the database of polymorphisms. For these four tongue tumor samples, we identified an average of 1474 total SVs per sample. Filtering these SVs using the Rare Variant Analysis pipeline for SVs not found in the Bionano control database yielded an average of 72 total SVs per sample, consisting of 11 insertions (range 9-15), 31 deletions (range 11-47), 3 inversions (range 1-6), 14 duplications (range 2-23), and 14 translocations (6-19). Filtering against the variants found in the corresponding blood samples returned an average of 58 total SVs per sample, consisting of 10 insertions (range 9-10), 20 deletions (range 7-35), 2 inversions (range 0-4), 13 duplications (range 4-24), and 14 translocations (range 6-19) (Figure 3, upper panel).

**Figure 3.**
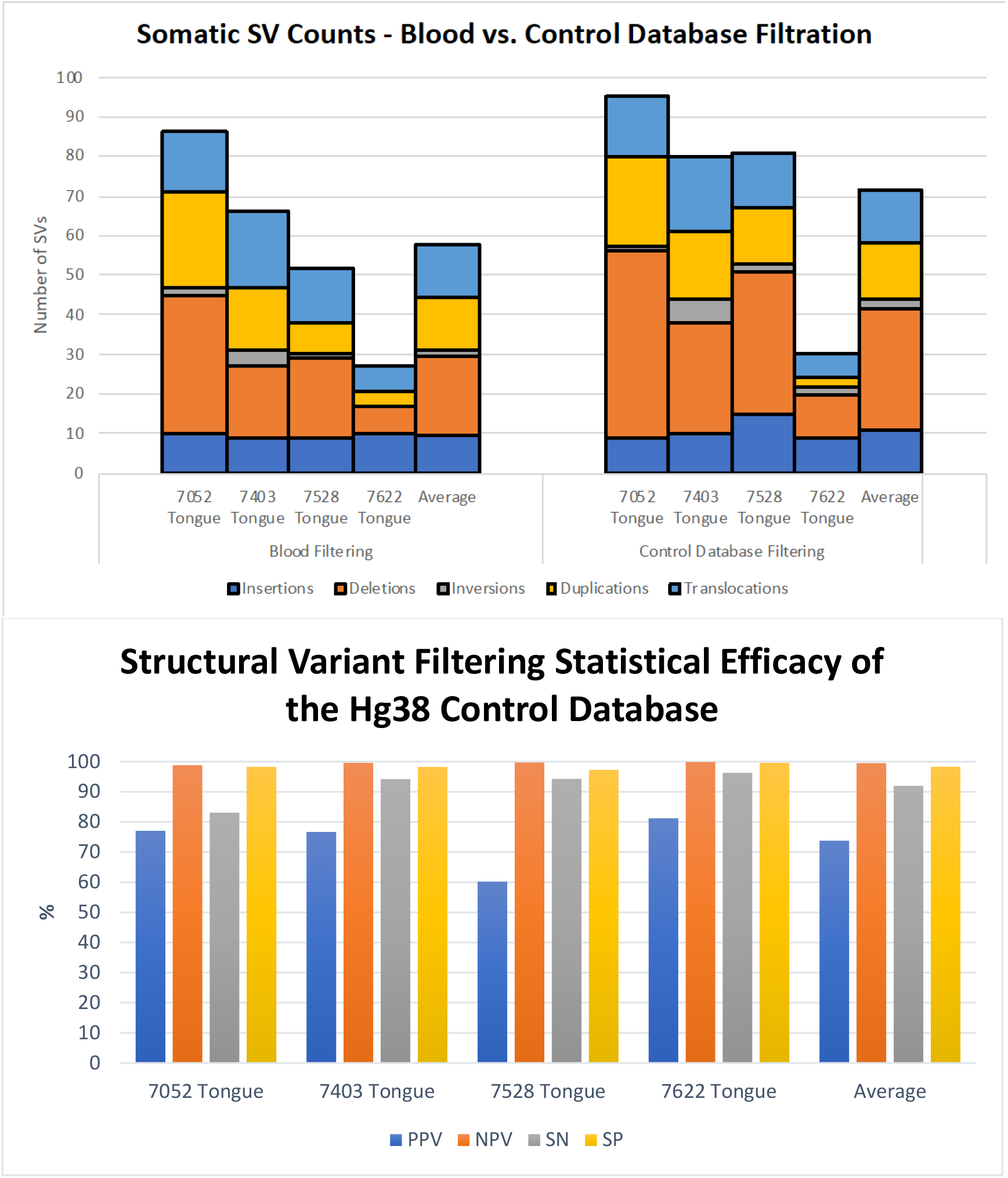
Efficacy of the Somatic Variant Identification Using a Control Database of Known Polymorphisms. Upper Panel: Number and distribution of somatic structural variant in four tongue tumors as determined by filtering against SVs in the patient’s genome from peripheral blood (left) or against Bionano’s control database of known polymorphisms. Lower Panel: Values for sensitivity, specificity and positive and negative predictive values for identification of somatic structural variants obtained by filtering total identified SVs to remove those present in a control database of know human polymorphisms. Data obtained by this filtering method were compared to those obtained by filtering the total SVs to remove those present in the genomes obtained from peripheral blood from the each of the patients from whom the tumors were removed.

Comparing the residual SV sets obtained by filtering against Bionano’s control database to the sets of true somatic SVs for each sample demonstrated that the control database filtration exhibited strong statistical accuracy (Figure 3, lower panel). Across the four separate samples, the control database exhibited an average sensitivity of 92% (83-96%) and specificity of 98% (range 97-99%). That is, filtering with the control database retained most of the true somatic mutations while eliminating almost all of the polymorphic SVs. Similarly, the average negative predictive value of the filter was 99.6%, demonstrating that an SV identified as germline was indeed a germline variant, while the positive predictive value of 74% (range 60-81%) indicates that a majority, but not all, the variants identified as somatic are in fact somatic. The SVs inaccurately identified as somatic were rare germline variants, predominantly insertions or deletions, essentially private to the patient’s genome. As above, we noted that the filtering process did not affect all SV types equally: while most deletions and insertions were flagged as polymorphic and eliminated from the list of somatic mutations, very few duplications and essentially no translocations were identified as polymorphic. This is consistent with observation that few translocations or duplications are stable through meiosis.

The number and types of somatic variants in a tumor varied substantially across the collection of samples (Figure 4). Several tumor samples, including those from colon, bladder, kidney and all four from thyroid, contained relatively few somatic SVs whereas others, including those from prostate, ovaries, lung and brain, carried a large number of somatic SVs. Since these samples for the most part serve as single representatives of each tumor type, we cannot extrapolate to the tumor types as a whole the contribution of SVs to cancer onset and development for each class of tumor. However, it is noteworthy that the SNV mutational burden in thyroid cancers is among the lowest among all tumor types and that measure of genome instability is mirrored in the low number of somatic SVs in all four of the samples examined [38]. Similarly, the SNV mutational burden in lung cancers is among the highest across all tumor types and both of the lung tumors examined here also carry a high level of somatic SV.

**Figure 4.**
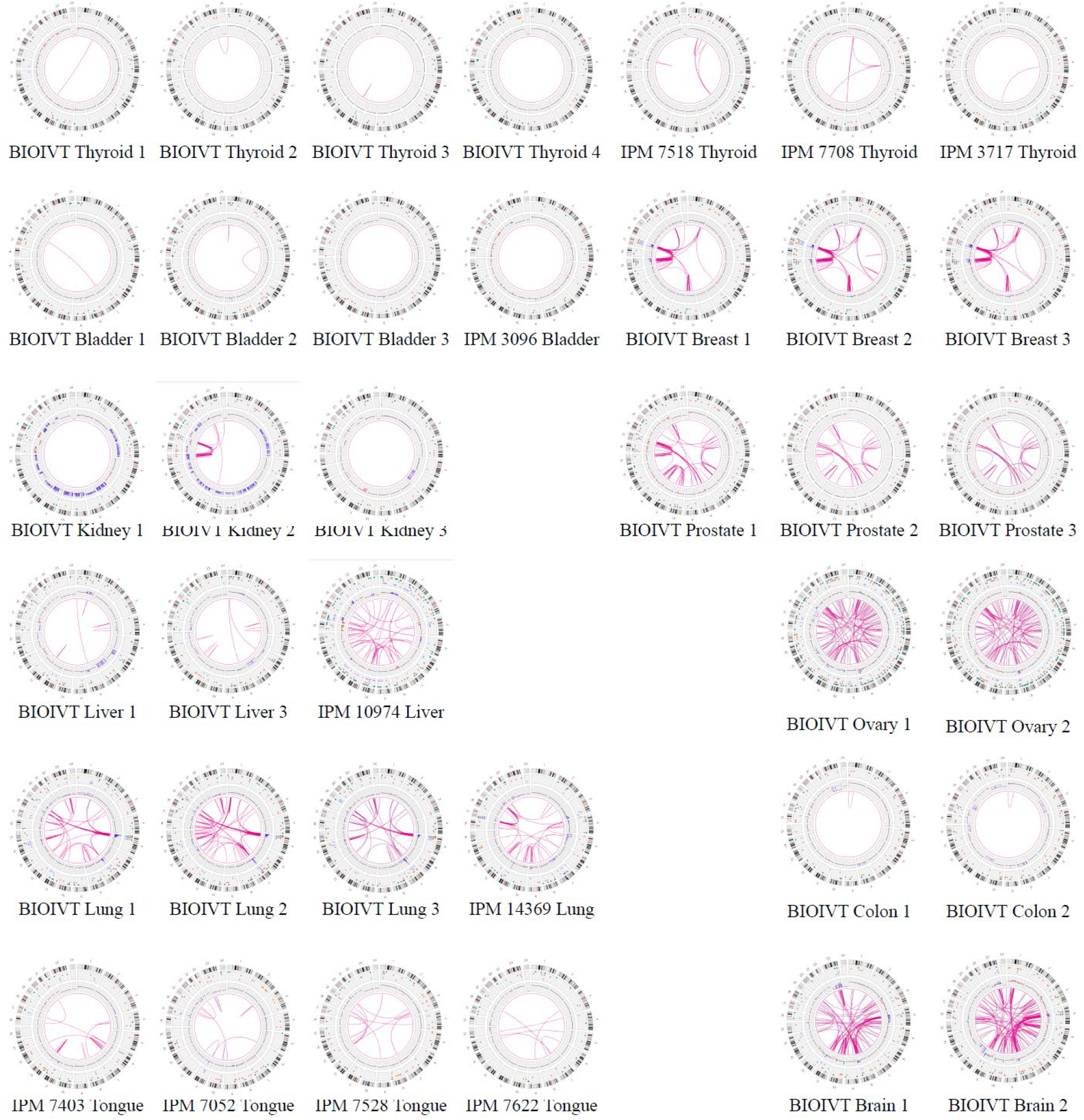
Global View of Structural Variants in Solid Tumor Samples. Diagrams of somatic structural variants in all the solid tumor genomes, filtered to remove known polymorphisms, showing translocations and inversions in the center, copy number on the inner ring and insertions (green), deletions (orange) inversions (light blue) and duplications (violet) on the next to most outer ring. Chromosomes are ordered sequentially in the outer ring on which are indicated cytological banding patterns and the centromere (red bar).

#### Duplicate Sample Analysis

We compared SV calls from separate isolates of the same sample to assess consistency and reproducibility of the method, albeit without knowing the extent of tumor heterogeneity of the individual samples. Six samples underwent triplicate analysis, and four samples underwent duplicate analysis (Table 2). After identifying SVs using the Rare Variant Analysis pipeline and filtering them against the Bionano control database of known polymorphisms, we recovered an average of 116 somatic SVs shared among the separate isolates of the same tumor. These comprised on average of 23 insertions, 29 deletions, 10 inversions, 11 duplications and 43 translocations (Table 3). As noted above, the number of SVs identified in a tumor varied widely across the different tumors examined, with lung, breast, brain and ovarian tumors showing a high level of somatic SVs while the others containing a relative low number of SVs. Moreover, the percentage of SVs shared among different isolates of the same tumor also varied among the different tumor types. However, the percentage of shared SVs and the total number of SVs were uncorrelated. Assuming that the higher values for shared SVs reflect the reproducibility of the method, then we might postulate that the lower shared values represent both the reproducibility and the tumor heterogeneity. This would suggest that these brain, liver, lung and prostate tumors had a relatively high level of tumor heterogeneity.

**Table 3.**
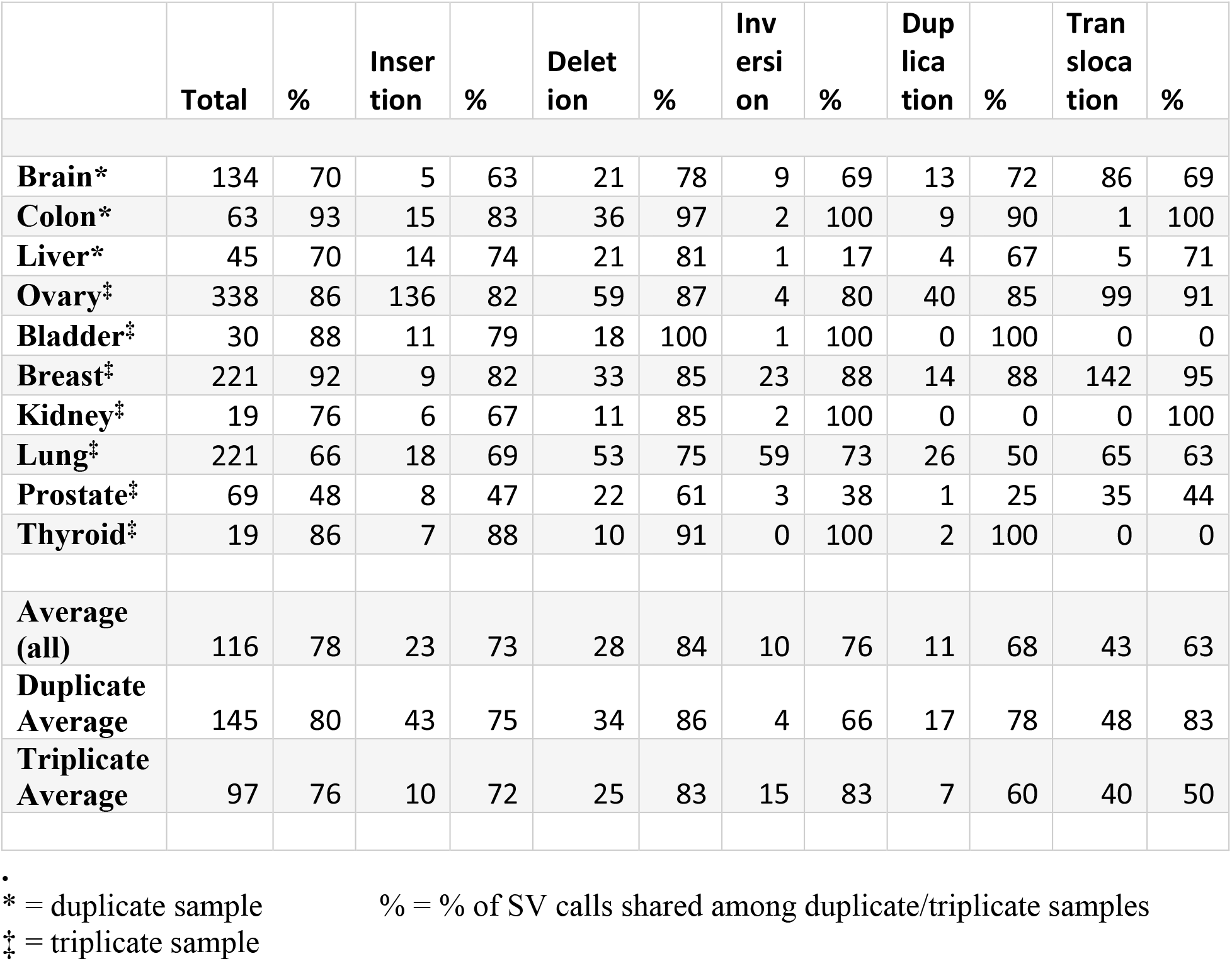
Duplicate Sample Analysis. Shown are the number of somatic structural variants shared among the multiple isolates of the same sample and the percentage of those relative the total number of somatic variants found in all the isolates of the same sample.

#### Identification of cancer gene mutations

While as noted above we cannot generalize regarding the role of structural variants in onset and progression of different tumor types, our results indicate that we can extract from the structural variant list clinically relevant data on individual tumors that might inform prognosis or treatment options. We examined the somatic structural variants in each tumor sample for those that affected genes previously associated with cancer. In particular, we annotated those genes altered by a structural variant, either by disruption, duplication, deletion or fusion, and intersected that list with the set of cancer-related genes in the Cosmic database (v92) [37]. The resultant list by tumor type is provided in Table 4 and subdivided into oncogenes, tumor suppressor genes and gene fusions. We included only those oncogenes that were potentially activated by duplication or gene fusion and only those tumor suppressor genes that were potentially inactivated by deletion, insertion or fusion. As evident, every tumor sample carried at least one such cancer gene mutation and most contained multiple hits. Several of these genes offer the opportunity for targeted therapies, focused either directly on the oncogene, as would be the case for CDK6 and ERBB2, or at the pathway downstream of the affected gene, as would be the case for BRAF and CDKN2A. Other affected genes, such as MSH2, RAD51B, RAD21 and RAD18, suggest that the potential of therapy based on possible ensuing genome instability, such as immunotherapy or PARP inhibitors. Many of these variants would not be readily identified by targeted gene panels generally used for clinical assessment of tumor genomes. Thus, the identification of somatic structural variants by OGM could provide useful clinical insights not readily available through standard next generation sequencing.

**Table 4.**
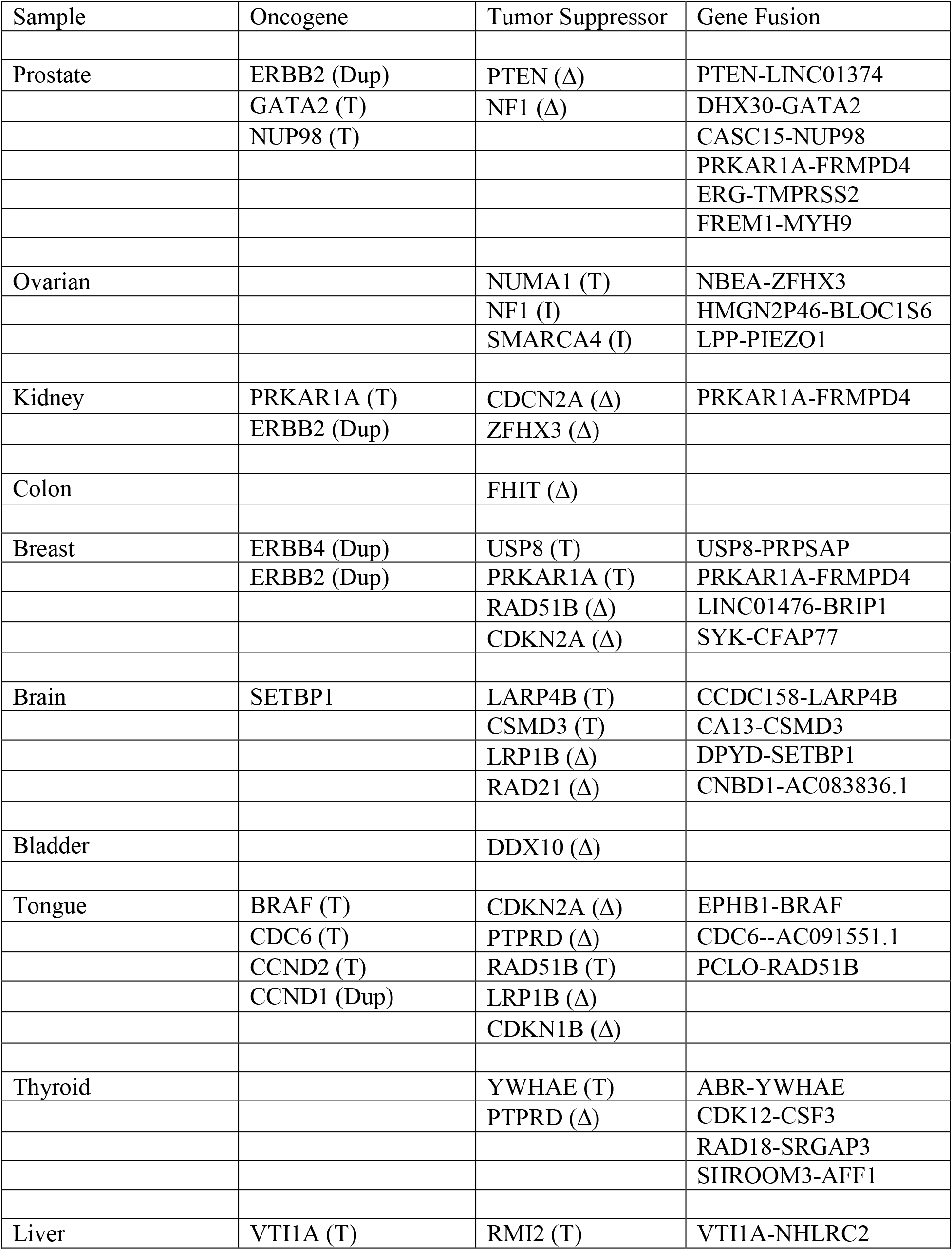
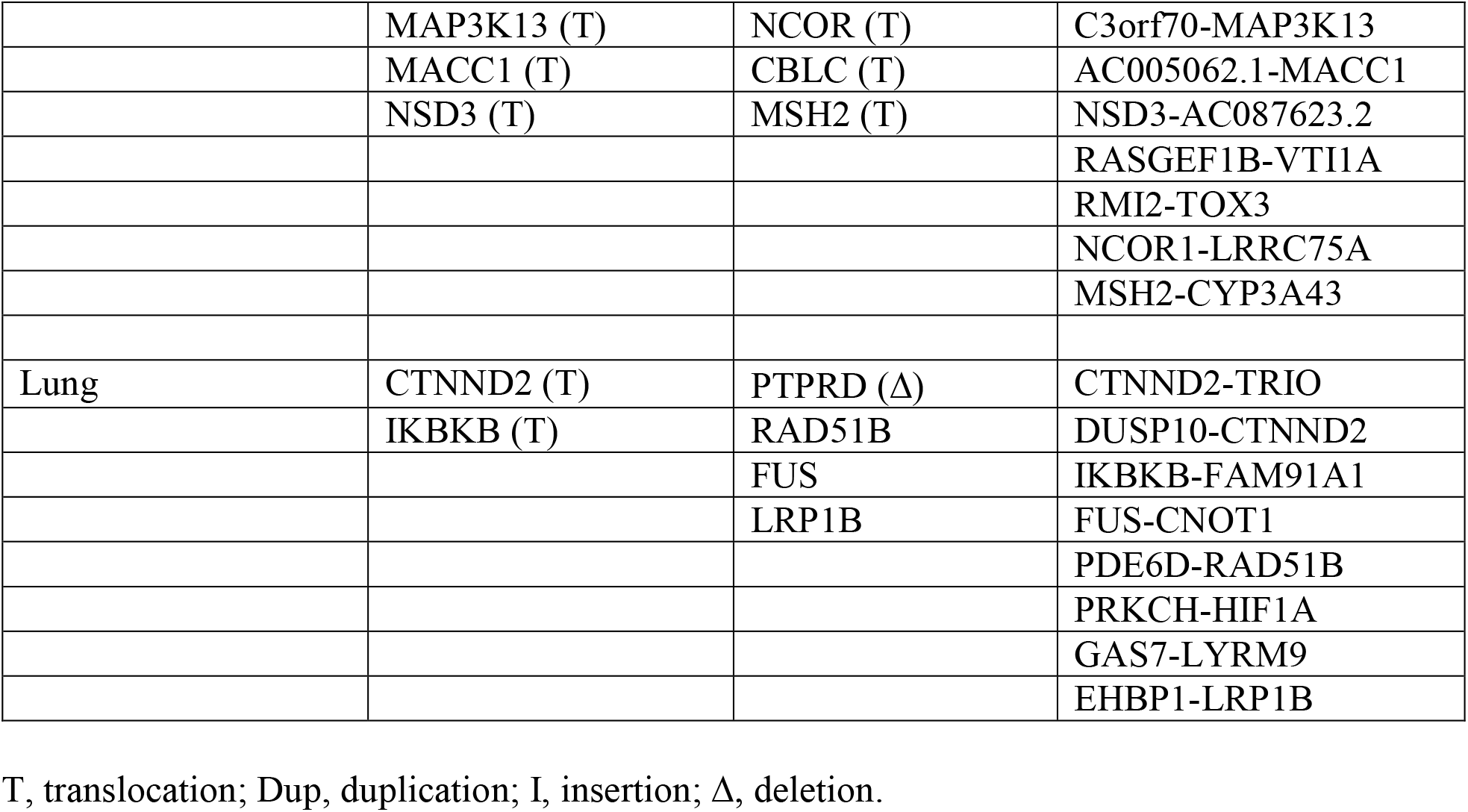
Structural Variants Affecting Cancer Relevant Genes.

In addition to identifying individual cancer related genes in tumor types, our results provide a panoramic view of the entire tumor genome and reveal large scale genomic features not readily available from standard sequencing techniques. As evident in the results in Figure 4, our data provide a rapid snapshot of the extent of genomic instability in each of the tumors. Such images present an integrated picture of the aneuploidies, translocations, inversions, deletions and insertions, which offers a readily digestible impression of the extent of genetic instability underlying a tumor. Moreover, several large-scale features are evident in these data. For instance, Figure 5 details a chromothrypsis event on a portion of chromosome 5 in one of the lung tumor samples. The detection and mapping of such a feature are difficult to achieve by short read sequencing but can indicate poor prognosis and the corresponding need for aggressive therapy.

**Figure 5.**
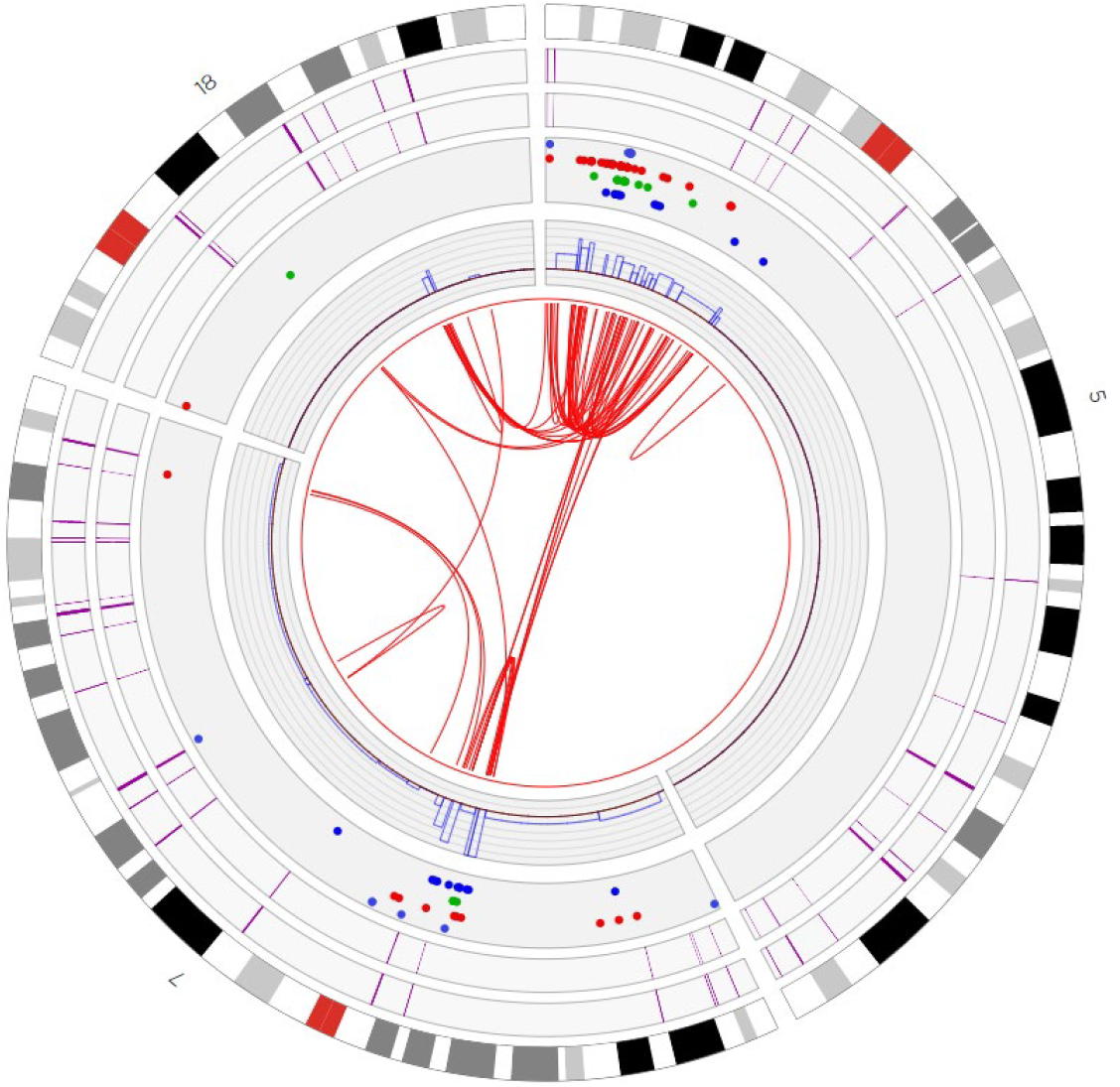
Chromothrypsis of Chromosome 5p in a Lung Tumor. Shown is a truncated Circos plot of the lung tumor, focused on the region of chromosome 5, highlighting the chromothrypsis event that occurred on its p arm. The organization of the circos plot is as indicated in the legend to Figure 4.

## Discussion

In this report we described the application of optical genome mapping to solid tumors, which we suggest can significantly augment the genomic analysis of such tumors obtained by next generation sequencing. Genomic analysis of tumors has stimulated major advances in cancer diagnosis, prognosis and treatment, shifting the focus from morphological and histochemical characterization to consideration of the landscape of driver mutations in the tumor [39-41].

Somatic driver events in a tumor – point mutations and structural variants (SVs) including insertions, deletions, inversions, translocations and copy number changes – are currently identified in solid tumors by some combination of RNA sequencing and genome sequencing of either targeted gene panels, whole exomes or whole genomes. As noted in this report, OGM can provide a pervasive view of the structural variants in a tumor and the cancer related genes on which they impinge, thus identifying affected genes agnostically, without prior bias imposed by gene panels.

In liquid tumors such as leukemia, genomic analysis is augmented by karyotyping, which gives a panoramic, albeit low resolution, view of the entire genome. Despite the low resolution, the genome wide view of the structural changes afforded by karyotyping reveals diagnostic features of the tumor that have strong prognostic value. Given the consistent correlation of clinical outcomes with specific mutation classes, the World Health Organization (WHO), National Comprehensive Cancer Network (NCCN) and European Leukemia Net (ELN) agencies developed recommendations for diagnosis and management of acute myeloid leukemia in adults based on the spectrum of somatic point mutations and SVs generally revealed by karyotyping. SVs, particularly translocations and inversions, are major considerations in this diagnosis. Since karyotyping is a very challenging technique to apply to solid tumors, the clinician does not have access to a comparable global view of a solid tumor’s genome and the role of SVs in prognosis has been possibly underappreciated. Applying OGM broadly to cancer types and correlating SVs revealed by that analysis could provide new genomics markers for prognosis and treatment selection.

Some prior studies have begun to demonstrate the utility of Bionano UHMW DNA isolation protocols in solid tissue tumor analysis. These include studies of lung squamous cell carcinoma and metastatic prostate carcinoma [7,34,35]. This current report demonstrates the utility of the a DNA isolation protocol and SV analysis in a wide variety of solid tissue types, and expands the feasibility of such analysis for previously unused human tissue types. The high DNA yield, high effective coverage, map rate and other molecular quality metrics shown across tumor types confirm how our extraction and analysis workflow can be effectively applied to many solid tissue tumors. This high-resolution genome imaging for many types of tumors will allow increased detection of cancer-associated SV events, potentially leading to new insights and improved personalized treatment options.

This current DNA isolation protocol carries a number of advantages. Tissue handling can be performed at room temperature. The current protocol showed successful DNA isolation in solid tissue samples of <20mg, and even as low as 6mg. The low tissue input requirement carries important applications for rare cancer samples, human tissue biopsy testing and other low-quantity specimen acquisition. Additionally, utilizing the novel paramagnetic Nanobind disks rather than prior agarose gel plugs greatly decreases time needed to complete DNA isolation to only 5 hours. The ability to isolate DNA from up to 8 simultaneous samples using the current protocol greatly amplifies throughput and reduces tissue-to-data processing time, increasing both laboratory convenience as well as expanding potential for clinical utility where rapid data turnaround is paramount. Furthermore, the strong inter-sample SV correspondence shown by most tissue types in duplicate/triplicate sample analysis demonstrates the reproducibility of this technique; intra-sample heterogeneity of select samples may be attributed to non-tumor normal tissue within some tissue fragments, or attributed to specific cancer subtype, and merits further investigation. Although the isolation protocol described here affords many advantages, there are some limitations to this protocol. While high quality DNA isolation and OGM SV analysis was obtained for a wide variety of tumor types that were tested, it may not be generalizable to every additional untested solid tumor type. Future directions include continuing to validate this protocol in additional tissue types, and assessing additional tumor samples to assess broader trends in the role of specific OGM-identified SVs in individual cancer subtypes.

## Conclusion

We demonstrate the utility of a DNA isolation protocol for high molecular weight DNA extraction and OGM SV analysis of a wide variety of solid human tumor types on the Bionano Saphyr system, including breast, colon, liver, brain, bladder, kidney, lung, ovary, prostate and thyroid cancer tissue. The system can be used to accurately detect genetic mutation hallmarks in cancer tissue samples, including rearrangements such as translocations, gene fusions and copy number alterations. Somatic SVs can be determined by comparison filtering with the Bionano control sample database, or against a matched pair sample. Importantly, Bionano SV pipelines can detect SVs with complex breakpoint structures that are difficult to detect with other technologies. Our results indicate that the solid tissue DNA extraction protocol can be applied to a wide variety of solid tumors, and the Saphyr system can capture in a streamlined workflow a broad spectrum of SVs with functional importance and provide great utility in cancer study.

## Data Availability

Data are available on request from James R. Broach

## Author Contributions

Conceptualization: DG, ARH, JRB; Methodology, HBS, HW, KP MO, YZ; Software, LZ; Validation, CYJL; Formal analysis, AWCP, BC, CYJL, DYG, LZ; Investigation, DYG, BL, SC; Writing - Initial Draft: DG, ARH; Writing – Editing: DG, AWCP, ARH; Writing - Final Draft: JRB; Visualization, DYG, ARH, BC; Supervision, ARH, DG, JRB, HBS, MO; Project administration, ARH, JRB; Funding acquisition, DG. All authors have read and agreed to the published version of the manuscript.

## Institutional Review Board

Patients were consented under protocol 40532 approved by the Penn State Health Institution Review Board

## Data Availability

Primary Bionano Saphyr data are available on request

